# Outcomes of Percutaneous Tracheostomy for Patients with SARS-CoV-2 Respiratory Failure

**DOI:** 10.1101/2021.02.23.21252231

**Authors:** Jason Arnold, Catherine A. Gao, Elizabeth Malsin, Kristy Todd, A. Christine Argento, Michael Cuttica, John M. Coleman, Richard G. Wunderink, Sean B. Smith, for the NU COVID Investigators

## Abstract

**Background:** SARS-CoV-2 can cause severe respiratory failure leading to prolonged mechanical ventilation. Data are just emerging about the practice and outcomes of tracheostomy in these patients. We reviewed our experience with tracheostomies for SARS-CoV-2 at our tertiary-care, urban teaching hospital.

**Methods:** We reviewed the demographics, comorbidities, timing of mechanical ventilation, tracheostomy, and ICU and hospital lengths-of-stay (LOS) in SARS-CoV-2 patients who received tracheostomies. Early tracheostomy was considered <14 days of ventilation. Medians with interquartile ranges (IQR) were calculated and compared with Wilcoxon rank sum, Spearman correlation, Kruskal-Wallis, and regression modeling.

**Results:** From March 2020 to January 2021, our center had 370 patients intubated for SARS-CoV-2, and 59 (16%) had percutaneous bedside tracheostomy. Median time from intubation to tracheostomy was 19 (IQR 17 – 24) days. Demographics and comorbidities were similar between early and late tracheostomy, but early tracheostomy was associated with shorter ICU LOS and a trend towards shorter ventilation. To date, 34 (58%) of patients have been decannulated, 17 (29%) before hospital discharge; median time to decannulation was 24 (IQR 19-38) days. Decannulated patients were younger (56 vs 69 years), and in regression analysis, pneumothorax was associated was associated with lower decannulation rates (OR 0.05, 95CI 0.01 – 0.37). No providers developed symptoms or tested positive for SARS-CoV-2.

**Conclusions:** Tracheostomy is a safe and reasonable procedure for patients with prolonged SARS-CoV-2 respiratory failure. We feel that tracheostomy enhances care for SARS-CoV-2 since early tracheostomy appears associated with shorter duration of critical care, and decannulation rates appear high for survivors.

## INTRODUCTION

Severe acute respiratory syndrome coronavirus 2 (SARS-CoV-2) infection may cause acute, severe respiratory failure that can lead to prolonged mechanical ventilation. At the onset of the pandemic there were questions regarding the safety and value of tracheostomy for SARS-CoV2 patients with prolonged respiratory failure. Tracheostomy has since been described and safely performed in these patients, and several societal guidelines support performing tracheostomy with proper personal protective equipment and precautions.^1-6^

Data are emerging about the practice and outcomes of tracheostomy for prolonged respiratory failure from SARS-CoV-2. Rates of decannulation and survival have varied in the literature. In the largest cohorts from Spain and England, median times to tracheostomy ranged from 12 to 16 days.^7,8^ The Spanish multicenter cohort found an overall mortality rate of 23%, and their decannulation rate for survivors weaned from ventilation was 81%.^7^ Another study from England reported 85% survival at 30 days and a 99% overall decannulation rate for survivors.^9^ Several smaller studies from earlier in the pandemic reported lower decannulation rates, ranging from 8% to 13%.^1,3,4^ However, one of the largest systematic review and meta-analysis comprising over 3000 patients found an average decannulation rate of 34.9.^7^ One potential explanation for differences is availability and willingness of long term acute care (LTAC) facilities to transfer SARS-CoV-2 infected patients.

We sought to review our practice at a large, US tertiary-care, urban teaching hospital that has had a high volume of patients with SARS-CoV-2 respiratory failure. Our goals were to determine mortality and decannulation rates as well to assess patient characteristics associated with successful outcomes.

## METHODS

We reviewed patients with SARS-CoV-2 who had percutaneous bedside tracheostomy for prolonged respiratory failure in our single-center, tertiary-care, urban teaching hospital from March 2020 to January 2021. These tracheostomies were bedside percutaneous procedures performed by interventional pulmonologists wearing powered air purifying respirators, gowns, and gloves. The procedures followed our standard practices for percutaneous tracheostomy, although the oropharynx was packed with gauze to minimize aerosolization when the cuff on the endotracheal tube was deflated. Patient demographics and comorbidities, the timing of mechanical ventilation and tracheostomy, as well as ICU and hospital lengths-of-stay (LOS) were catalogued. Primary outcomes included overall mortality and decannulation rates, whereas the secondary outcome was time to weaning from mechanical ventilation. The timing of tracheostomy was at the discretion of the ICU attending and interventional pulmonologist who performed the procedures. Tracheostomy was considered early when performed within 14 days of initiation of mechanical ventilation. Like other centers, we experienced two waves of admissions: we considered the first wave those admitted from March to July 2020; whereas the second wave included those admitted after August 1, 2020.

Statistical analyses were performed with Stata 11.2 (College Station, TX). Not all continuous data were normally distributed, and so median values with interquartile ranges (IQR) were calculated. Non-parametric analyses included Wilcoxon rank sum and Spearman correlation testing. Kruskal-Wallis testing was used to compare data across multiple categories. Regression modeling was used to identify variables associated with outcomes. Statistical significance was established as p < 0.05.

## RESULTS

From March 2020 to January 2021, 370 patients were intubated for SARS-CoV-2 respiratory failure, and percutaneous bedside tracheostomy was performed in 59 (15.9%) patients. Median age was 66 (IQR 61 – 71) years, and 32% were female. Median body mass index was 27 (24 – 33), and the median number of comorbidities was 2 (IQR 1 – 3). The most common comorbidities were hypertension (61%), diabetes (54%), and obesity (41%). Twenty-six patients (44%) were treated with steroids, and 21 (36%) with remdesivir, either under the emergency use authorization or following FDA approval. An additional 5 (8%) patients participated in a double blind remdesivir versus placebo trial.

Median time from intubation to tracheostomy was 19 (IQR 17 – 24) days. Median PEEP was 10 (IQR 6 – 10), and FiO2 was 40% (40% – 50%) on the day of tracheostomy. The most common tracheostomy placed was a Shiley 6 distal XLT (n=36, 61%). No procedural complications related to tracheostomy placement occurred. The most common overall hospital complications were pneumonia (83%), and the majority of pneumonia cases (37/49) occurred before tracheostomy placement. Venous thromboembolism (56%), acute renal failure requiring dialysis (44%), and pneumothorax (24%) were the other common hospital complications.

Median length of follow-up has been 42 (IQR 16 – 125) days, and 81% have 30-day follow-up data available. Decannulation occurred in 34 patients (83% of survivors and 58% of all patients), with 17 patients (50%) being decannulated prior to hospital discharge. Median time to decannulation was 24 (IQR 19 – 38) days. Median ICU LOS was 37 (IQR 32 – 43) days, and hospital LOS was 41 (35 – 54) days. Median duration of mechanical ventilation was 35 (IQR 32 – 41) days, and median time from tracheostomy to weaning from mechanical ventilation was 17 (10 – 19) days.

Overall mortality was 32%, and hospital mortality was 29%. Only acute renal failure requiring dialysis (OR 3.18, 95CI 1.02 – 9.92, p=0.046) and pneumothorax (OR 6.30, 1.72 – 23.11, p=0.006) were associated with overall mortality. Patients who died had a higher FiO2 at the time of tracheostomy (50 [IQR 40 – 50] vs. 40 [40 – 50], p=0.043), although FiO2 >40% was not associated with mortality (OR 2.57, 95CI 0.83 – 7.93, p=0.100).

Demographics and comorbidities were similar for those who had tracheotomy before or after 14 days of mechanical ventilation, but the FiO2 at the time of tracheotomy was lower in those who had early tracheostomy (40 [IQR 40 – 50] vs 50 [40 – 50], p=0.037). Early tracheostomy was associated with shorter ICU duration (30 [IQR 27 – 38] vs. 38 [34 – 44] days, p=0.017), and a trend towards shorter duration of mechanical ventilation (34 [24 – 35] vs 35 [33 – 44] days, p=0.056). Neither survival nor decannulation rates differed between early or late tracheostomy.

Patients who were decannulated were younger (56 [IQR 37 – 63] vs. 69 [65 – 72] years, (p<0.001), had higher BMI (30 [24 – 35] vs. 26 [24 – 28], p=0.037), and lower FiO2 at the time of tracheostomy (40 [IQR 40 – 50] vs. 50 [40 – 50], p=0.037). There was a trend towards shorter time from intubation to tracheostomy (19 [IQR 14 – 22] vs. 21 [18 – 26] days, p=0.050) for those who were decannulated. Four univariable characteristics were associated with decannulation: steroids (OR 0.32, 0.11 – 0.93, p=0.037); remdesivir (OR 0.20, 0.06 – 0.64, p=0.007); pneumothorax (OR 0.07, 0.01 – 0.34, p=0.001); obesity (OR 3.56, 1.14 – 11.12, p=0.029). When these four variables were analyzed in multiple regression, only pneumothorax remained significantly associated with lower decannulation rates (OR 0.05, 95CI 0.01 – 0.37, p=0.004).

To date, 36 survivors (64.3%) were weaned from mechanical ventilation by hospital discharge. Demographics, comorbidities, and tracheostomy characteristics were not associated with time to weaning. Female gender was associated with increased likelihood of being weaned from the ventilator by discharge (OR 4.53, 95CI 1.13 – 18.24, p=0.033), whereas acute renal failure was associated a lower rate of weaning by discharge (OR 0.27, 95CI 0.09 – 0.85, p=0.025). There was a non-significant trend towards shorter weaning in patients who had received steroids (15 [IQR 8 – 19] vs 19 [12 – 20] days, p=0.064).

No providers involved in the placement of tracheostomies developed symptoms or tested positive for SARS-CoV-2.

### Subgroup analysis by admission date

There were 45 patients who had tracheostomy in the first wave and 14 in the second. In the second wave, 3 of the 14 patients remained admitted at the time of this study. Baseline demographics (i.e., age, gender, comorbidities) and times from intubation to tracheostomy were similar between the two waves. There was significantly higher utilization of steroids (92.9% vs. 28.9%, p<0.001) and remdesivir (85.7% vs. 20.0%, p<0.001) in the second wave. Rates of renal failure and pneumonia were similar, but there were trends toward higher rates of pneumothorax (42.9% vs. 17.8%, p=0.056) and venous thromboemboli (78.6% vs. 48.9%, p=0.053) in the second wave. Mortality, hospital LOS, ICU LOS, duration of mechanical ventilation, and time to weaning from mechanical ventilation were similar between the two waves. Decannulation by hospital discharge was higher in the first wave (37.8% vs 0%, p=0.016).

## DISCUSSION

We found that tracheostomy for SARS-CoV-2 patients was a safe and reasonable practice for prolonged respiratory failure. As described in similar studies, we found no incidents of operators contracting SARS-CoV-2 infection during tracheostomy.^2,5,6,8-10^

The ideal time from intubation to tracheostomy for SARS-CoV-2 has been debated, as it has been for other critical illnesses. There had been initial concerns that tracheostomy should be delayed until after active SARS-CoV-2 viral replication, whereas others proposed early tracheostomy to facilitate weaning and preserve resources during pandemic.^11^ Our practice has been to maintain traditional standards for tracheostomy selection with regard to timing, oxygenation, and hemodynamic stability. Although we do not have data regarding sedation dosing before and after tracheostomy, we suspect that tracheostomy facilitates lightening sedation and weaning SARS-CoV-2 patients as in other causes of prolonged respiratory failure.^7^

Our median times to tracheostomy (19 days) and times to weaning (17 days) are similar to other reports in the literature.^2-4,12,13^ Mata-Castro et al. (2021) found that a longer time from intubation to tracheostomy was related to a longer time from tracheostomy to weaning.^13^ Two other studies found that early tracheostomy was associated with shorter overall duration of mechanical ventilation and ICU LOS.^2,6^ We did not find an association between the time to tracheostomy and the outcomes of weaning, decannulation, or mortality, although there was shorter ICU LOS after early tracheostomy.

Differences between the initial surge in the spring of 2020 and a second wave later in the fall have been described. Subgroup analyses between our two waves found trends towards more pneumothorax and venous thromboemboli in the second wave, and undoubtedly more patients received steroids in the second wave because of the RECOVERY trial.^14^

Studies have demonstrated overall relatively good survival rates for patients with SARS-CoV-2 who had tracheostomy, with mortality ranging from 7% to 23%.^1,3,5^ Many studies have been affected by duration of follow-up and available outcome data, and the largest study from Spain reports one of the higher mortality rates of 23%.^7^ Complete 30-day follow-up data were available for 81% of our patients, and this subgroup had a 33% 30-day mortality rate. This elevation in mortality found in our study may be attributed to the high number of external hospital transfers (n=9, 15%) for escalation of care. Survival amongst tracheostomy patients has been described as higher than those who did not receive tracheostomy,^9^ although selection bias is to be considered for which patients are stable enough to have tracheostomy.

Long-term outcomes of lung function are unknown for patients who survive SARS-CoV-2 respiratory failure. Short-term decannulation rates for non-SARS-CoV-2 ARDS are not well published, but already multiple studies, including ours, have demonstrated high rates of decannulation within only a few months after ARDS from SARS-CoV-2. More than 80% of our surviving patients have been decannulated, although long-term lung function is yet unknown. We have used our data to counsel families when deciding upon tracheostomy, as we feel that tracheostomy is a safe and appropriate procedure that can facilitate weaning from mechanical ventilation.

## Supporting information

Appendix 1: NU COVID Investigators

## Data Availability

Data are available upon reasonable request.

## Acknowledgements

The authors thank the many individuals (nurses, respiratory therapists, social workers, physical therapists, and providers) who took care of the patients with COVID-19.

## TABLES

**Table 1:**
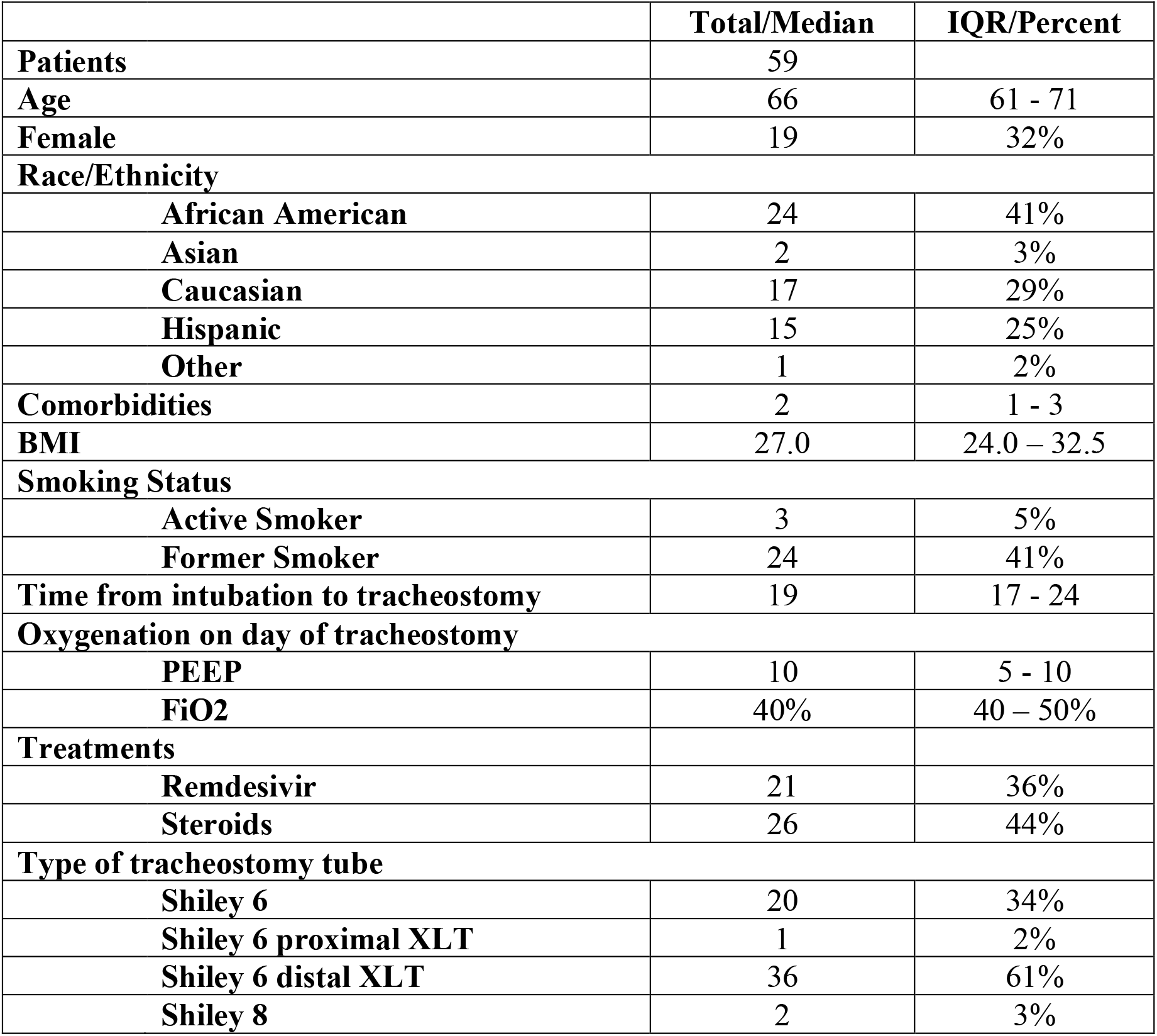
Baseline characteristics of patients with SARS-CoV-2 who had tracheostomy.

|  | **Total/Median** | **IQR/Percent** |
| --- | --- | --- |
| **Patients** | 59 |  |
| **Age** | 66 | 61 - 71 |
| **Female** | 19 | 32% |
| **Race/Ethnicity** |  |  |
| **African American** | 24 | 41% |
| **Asian** | 2 | 3% |
| **Caucasian** | 17 | 29% |
| **Hispanic** | 15 | 25% |
| **Other** | 1 | 2% |
| **Comorbidities** | 2 | 1 - 3 |
| **BMI** | 27.0 | 24.0 – 32.5 |
| **Smoking Status** |  |  |
| **Active Smoker** | 3 | 5% |
| **Former Smoker** | 24 | 41% |
| **Time from intubation to tracheostomy** | 19 | 17 - 24 |
| **Oxygenation on day of tracheostomy** |  |  |
| **PEEP** | 10 | 5 - 10 |
| **FiO2** | 40% | 40 – 50% |
| **Treatments** |  |  |
| **Remdesivir** | 21 | 36% |
| **Steroids** | 26 | 44% |
| **Type of tracheostomy tube** |  |  |
| **Shiley 6** | 20 | 34% |
| **Shiley 6 proximal XLT** | 1 | 2% |
| **Shiley 6 distal XLT** | 36 | 61% |
| **Shiley 8** | 2 | 3% |

**Table 2:** Comorbidities of patients with SARS-CoV-2 who had tracheostomy.

| **Comorbidities** | **N** | **Percent** |
| --- | --- | --- |
| Hypertension | 36 | 61% |
| Diabetes Mellitus | 32 | 54% |
| Obesity | 24 | 41% |
| Coronary Artery Disease | 10 | 17% |
| Chronic Kidney Disease | 10 | 17% |
| Obstructive Sleep Apnea | 8 | 14% |
| Solid Organ Malignancy | 9 | 15% |
| Heart Failure | 7 | 12% |
| Cerebrovascular Disease | 5 | 8% |
| Atrial Fibrillation | 5 | 8% |
| Chronic Lung Disease | 4 | 7% |
| Solid Organ Transplantation | 4 | 7% |
| End Stage Renal Disease | 2 | 3% |
| Deep Vein Thrombosis / Pulmonary Embolism | 2 | 3% |
| Cirrhosis | 2 | 3% |
| Hematologic Malignancy | 2 | 3% |

**Table 3:** Hospital complications of patients with SARS-CoV-2 who had tracheostomy.

| **Complications** | **N** | **Percent** |
| --- | --- | --- |
| Pneumonia | 49 | 83% |
| Pneumonia prior to tracheostomy | 37 | 63% |
| Deep vein thrombosis / pulmonary embolism | 33 | 56% |
| Acute Renal Failure requiring hemodialysis | 26 | 44% |
| Pneumothorax | 14 | 24% |
| Atrial or ventricular arrhythmia | 7 | 12% |
| Bacteremia | 7 | 12% |
| Hemorrhage | 5 | 8% |
| Acute Stroke | 5 | 8% |
| Cardiomyopathy | 4 | 7% |
| Cardiac arrest | 2 | 3% |
| Diabetic ketoacidosis | 2 | 3% |
| Rhabdomyolysis | 2 | 3% |
| Tracheal stenosis | 1 | 2% |
| Cardiac tamponade | 1 | 2% |

**Table 4:**
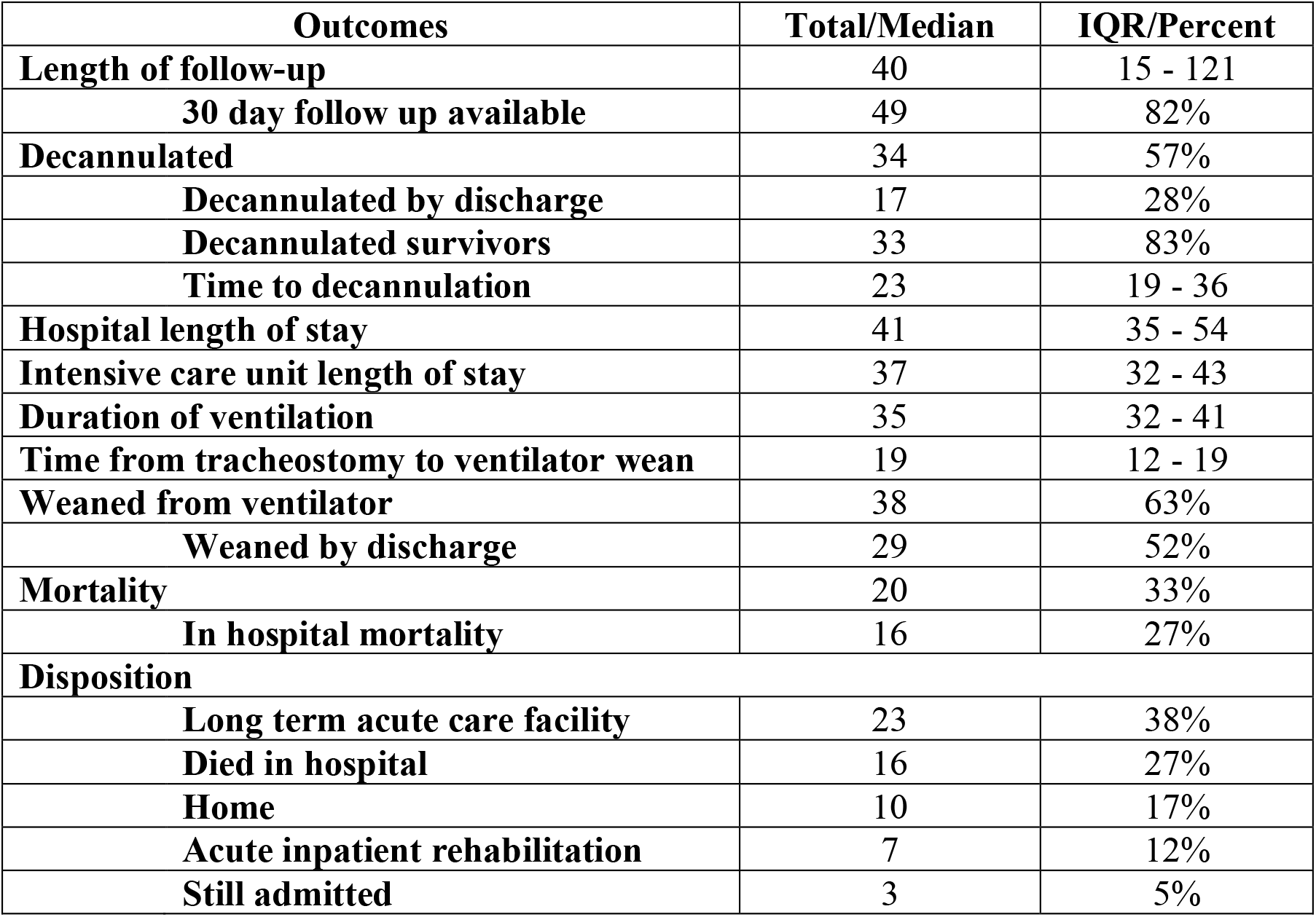
Outcomes of patients with SARS-CoV-2 who had tracheostomy.

| **Outcomes** | **Total/Median** | **IQR/Percent** |
| --- | --- | --- |
| **Length of follow-up** | 40 | 15 - 121 |
| **30 day follow up available** | 49 | 82% |
| **Decannulated** | 34 | 57% |
| **Decannulated by discharge** | 17 | 28% |
| **Decannulated survivors** | 33 | 83% |
| **Time to decannulation** | 23 | 19 - 36 |
| **Hospital length of stay** | 41 | 35 - 54 |
| **Intensive care unit length of stay** | 37 | 32 - 43 |
| **Duration of ventilation** | 35 | 32 - 41 |
| **Time from tracheostomy to ventilator wean** | 19 | 12 - 19 |
| **Weaned from ventilator** | 38 | 63% |
| **Weaned by discharge** | 29 | 52% |
| **Mortality** | 20 | 33% |
| **In hospital mortality** | 16 | 27% |
| **Disposition** |  |  |
| **Long term acute care facility** | 23 | 38% |
| **Died in hospital** | 16 | 27% |
| **Home** | 10 | 17% |
| **Acute inpatient rehabilitation** | 7 | 12% |
| **Still admitted** | 3 | 5% |

**Table 5:** Subgroup analysis comparing early tracheostomy (trach placement prior to 14 days of intubation) with late (>14 days of intubation) tracheostomy placement.

| **Outcomes** | **Early Tracheostomy** | **Late Tracheostomy** | **P value** |
| --- | --- | --- | --- |
| **Weaned from ventilator by discharge** | 50% | 49% | p = 0.44 |
| **Duration of mechanical ventilation** | 34 (27-34) | 36 (33-45) | p = 0.055 |
| **Decannulated** | 75% | 53% | p = 0.30 |
| **Hospital length of stay** | 35 (33-45) | 42 (37 – 54) | p = 0.14 |
| **Intensive care unit length of stay** | 30 (27 – 37) | 39 (35 – 44) | p = 0.016 |
| **Mortality** | 17% | 40% | p = 0.23 |
| **In hospital mortality** | 17% | 32% | p = 0.65 |

