## Appendix 1: NU COVID Investigators for "Outcomes of Percutaneous Tracheostomy for Patients with SARS-CoV-2 Respiratory Failure"

| NU COVID Investigators | ORCID | email address | Affiliation 1 |
|---|---|---|---|
| A. Christine Argento | 0000-0002-3807-7690 | | Pulmonary and Critical Care Medicine |
| Ajay A. Wagh | 0000-0003-3402-0272 | | Pulmonary and Critical Care Medicine |
| Alexandra C. McQuattie-Pimental | 0000-0003-4028-9905 | | Pulmonary and Critical Care Medicine |
| Alexander V. Misharin | 0000-0003-2879-3789 | | Pulmonary and Critical Care Medicine |
| Alexis Rose Wolfe | 0000-0001-9321-892X | | Pulmonary and Critical Care Medicine |
| Alvaro Donayre | 0000-0003-1877-0655 | | Pulmonary and Critical Care Medicine |
| Ankit Bharat | 0000-0002-1248-0457 | | Thoracic Surgery |
| Anna E. Pawlowski | 0000-0002-9047-6437 | | Northwestern Medicine Enterprise Data Warehouse |
| Anne R. Levenson | 0000-0002-7587-1885 | | Pulmonary and Critical Care Medicine |
| Anthony M. Joudi | 0000-0003-1232-3256 | | Pulmonary and Critical Care Medicine |
| Benjamin D. Singer | 0000-0001-5775-8427 | | Pulmonary and Critical Care Medicine |
| Betty Tran | 0000-0001-6794-7088 | | Pulmonary and Critical Care Medicine |
| Catherine A. Gao | 0000-0001-5576-3943 | | Pulmonary and Critical Care Medicine |
| Chao Qi | 0000-0003-0741-3070 | | Department of Pathology |
| Chiagozie O. Pickens | 0000-0002-3400-9228 | | Pulmonary and Critical Care Medicine |
| Chitaru Kurihara | 0000-0003-3536-4675 | | Thoracic Surgery |
| Clara J Schroedl | 0000-0003-3280-4997 | | Pulmonary and Critical Care Medicine |
| Daniel Meza | 0000-0001-8154-1295 | | Pulmonary and Critical Care Medicine |
| Daniel Schneider | 0000-0002-3523-745X | | Northwestern Medicine Enterprise Data Warehouse |
| David A. Kidd | 0000-0003-0411-9511 | | Pulmonary and Critical Care Medicine |
| David D. Odell | 0000-0002-0121-4831 | | Thoracic Surgery |
| David W. Kamp | 0000-0001-5796-1206 | | Pulmonary and Critical Care Medicine |
| Elizabeth S. Malsin | 0000-0002-3294-6752 | | Pulmonary and Critical Care Medicine |
| Emily M. Leibenguth | 0000-0001-7809-7947 | | Pulmonary and Critical Care Medicine |
| Eric P. Cantey | 0000-0002-7065-7169 | | Cardiology |
| Gabrielle Y. Liu | 0000-0003-2049-423 | | Pulmonary and Critical Care Medicine |
| GR Scott Budinger | 0000-0002-3114-5208 | | Pulmonary and Critical Care Medicine |
| Helen K. Donnelly | 0000-0001-8827-8581 | | Pulmonary and Critical Care Medicine |
| Isaac A. Goldberg | 0000-0002-7178-9729 | | Pulmonary and Critical Care Medicine |
| Jacob I. Sznajder | 0000-0002-2089-4764 | | Pulmonary and Critical Care Medicine |
| Jacqueline M. Kruser | 0000-0003-3258-1869 | | Pulmonary and Critical Care Medicine |
| James M. Walter | 0000-0001-7428-3101 | | Pulmonary and Critical Care Medicine |
| Jane E. Dematte | 0000-0003-4907-9525 | | Pulmonary and Critical Care Medicine |
| Jason M. Arnold | 0000-0001-8687-0063 | | Department of Medicine |
| John Coleman | 0000-0003-1246-3814 | | Pulmonary and Critical Care Medicine |
| Joseph Isaac Bailey | 0000-0002-4225-3108 | | Pulmonary and Critical Care Medicine |
| Joseph S. Deters | 0000-0002-3522-3824 | | Pulmonary and Critical Care Medicine |
| Justin A. Fiala | 0000-0001-5196-7076 | | Pulmonary and Critical Care Medicine |

| | | | |
|---|---|---|---|
| Katharine Secunda | 0000-0003-0310-5783 | | Pulmonary and Critical Care Medicine |
| Kaitlyn Vitale | 0000-0002-8679-6200 | | Pulmonary and Critical Care Medicine |
| Khalilah L. Gates | 0000-0002-5965-8136 | | Pulmonary and Critical Care Medicine |
| Kristy Todd | 0000-0001-8210-8522 | | Pulmonary and Critical Care Medicine |
| Lindsey D. Gradone | 0000-0001-8145-2997 | | Pulmonary and Critical Care Medicine |
| Lindsey N. Textor | 0000-0001-7699-9429 | | Pulmonary and Critical Care Medicine |
| Lisa F. Wolfe | 0000-0001-9044-4147 | | Pulmonary and Critical Care Medicine |
| Lorenzo L. Pesce | 0000-0002-8015-4653 | | Thoracic Surgery |
| Luisa Morales-Nebreda | 0000-0002-7126-0518 | | Pulmonary and Critical Care Medicine |
| Madeline L Rosenbaum | 0000-0001-9866-9850 | | Pulmonary and Critical Care Medicine |
| Manu Jain | 0000-0003-1534-5629 | | Pulmonary and Critical Care Medicine |
| Marc A. Sala | 0000-0002-2900-0595 | | Pulmonary and Critical Care Medicine |
| Mary Carns | 0000-0002-5063-156X | | Pulmonary and Critical Care Medicine |
| Marysa V. Leyla | 0000-0002-6583-5389 | | Cardiology |
| Mengjia Kang | 0000-0002-1679-9473 | | Pulmonary and Critical Care Medicine |
| Michael J. Alexander | 0000-0001-6271-3426 | | Pulmonary and Critical Care Medicine |
| Michael J. Cuttica | 0000-0002-0084-8760 | | Pulmonary and Critical Care Medicine |
| Michelle Hinsch Prickett | 0000-0002-1119-0079 | | Pulmonary and Critical Care Medicine |
| Natalie Jensma | 0000-0002-2046-5070 | | Hospital Medicine |
| Nicole Borkowski | 0000-0001-8650-6456 | | Pulmonary and Critical Care Medicine |
| Nikolay S. Markov | 0000-0002-3659-4387 | | Pulmonary and Critical Care Medicine |
| Orlyn R. Rivas | 0000-0001-6938-4020 | | Department of Medicine |
| Paul A. Reyfman | 0000-0002-6435-6001 | | Pulmonary and Critical Care Medicine |
| Peter H. S. Sporn | 0000-0002-1006-9437 | | Pulmonary and Critical Care Medicine |
| Prasanth Nannapaneni | 0000-0001-8080-3214 | | Northwestern Medicine Enterprise Data Warehouse |
| Rachel B. Kadar | 0000-0002-2583-3637 | | Critical Care Medicine, Anesthesiology |
| Rachel M. Kaplan | 0000-0003-0038-9231 | | Cardiology |
| Rade Tomic | 0000-0003-2394-1287 | | Pulmonary and Critical Care Medicine |
| Radhika Patel | 0000-0002-1255-5818 | | Pulmonary and Critical Care Medicine |
| Rafael Garza-Castillon | 0000-0003-4970-6584 | | Thoracic Surgery |
| Ravi Kalhan | 0000-0003-2443-0876 | | Pulmonary and Critical Care Medicine |
| Richard G. Wunderink | 0000-0002-8527-4195 | | Pulmonary and Critical Care Medicine |
| Rogan A. Grant | 0000-0003-0655-0882 | | Pulmonary and Critical Care Medicine |
| Romy Lawrence | 0000-0002-6578-8519 | | Pulmonary and Critical Care Medicine |
| Ruben J. Mylvaganam | 0000-0002-7203-7609 | | Pulmonary and Critical Care Medicine |

| | | | |
|---|---|---|---|
| Sean B. Smith | 0000-0001-8796-3966 | | Pulmonary and Critical Care Medicine |
| Samuel S. Kim | 0000-0001-8889-478X | | Thoracic Surgery |
| Sanket Thakkar | 0000-0002-0026-6533 | | Thoracic Surgery |
| SeungHye Han | 0000-0001-5625-6337 | | Pulmonary and Critical Care Medicine |
| Sharon R. Rosenberg | 0000-0002-0260-2545 | | Pulmonary and Critical Care Medicine |
| Susan R. Russell | 0000-0003-3006-1018 | | Pulmonary and Critical Care Medicine |
| Sydney M. Hyder | 0000-0002-6395-9062 | | Pulmonary and Critical Care Medicine |
| Taylor A. Poor | 0000-0002-2191-5037 | | Pulmonary and Critical Care Medicine |
| Theresa A. Lombardo | 0000-0002-7468-7541 | | Pulmonary and Critical Care Medicine |
| Zasu M. Klug | 0000-0003-3483-396X | | Pulmonary and Critical Care Medicine |

Table 1. NU COVID Investigators
